# Outpatient treatment with concomitant vaccine-boosted convalescent plasma for patients with immunosuppression and COVID-19

**DOI:** 10.1101/2023.08.29.23293790

**Authors:** Juan G. Ripoll, Sidna M. Tulledge-Scheitel, Anthony A. Stephenson, Shane Ford, Marsha L. Pike, Ellen K. Gorman, Sara N. Hanson, Justin E. Juskewitch, Alex J. Miller, Solomiia Zaremba, Erik A. Ovrom, Raymund R. Razonable, Ravindra Ganesh, Ryan T. Hurt, Erin N. Fischer, Amber N. Derr, Michele R. Eberle, Jennifer J. Larsen, Christina M. Carney, Elitza S. Theel, Sameer A. Parikh, Neil E. Kay, Michael J. Joyner, Jonathon W. Senefeld

**Author notes:** **Correspondence:** Jonathon W. Senefeld, PhD, Department of Health and Kinesiology, University of Illinois Urbana-Champaign | 906 S Goodwin | Urbana, Illinois 61801 |. Dr. Ripoll and Dr. Tulledge-Scheitel contributed equally as first authors to the work of the study and manuscript. Drs. Joyner and Senefeld contributed equally as senior authors to the work of the study and manuscript.

## Abstract

Although severe coronavirus disease 2019 (COVID-19) and hospitalization associated with COVID-19 are generally preventable among healthy vaccine recipients, patients with immunosuppression have poor immunogenic responses to COVID-19 vaccines and remain at high risk of infection with SARS-CoV-2 and hospitalization. Additionally, monoclonal antibody therapy is limited by the emergence of novel SARS-CoV-2 variants that have serially escaped neutralization. In this context, there is interest in understanding the clinical benefit associated with COVID-19 convalescent plasma collected from persons who have been both naturally infected with SARS-CoV-2 and vaccinated against SARS-CoV-2 (“vax-plasma”). Thus, we report the clinical outcome of 386 immunocompromised outpatients who were diagnosed with COVID-19 and who received contemporary COVID-19 specific therapeutics (standard of care group) and a subgroup who also received concomitant treatment with very high titer COVID-19 convalescent plasma (vax-plasma group) with a specific focus on hospitalization rates. The overall hospitalization rate was 2.2% (5 of 225 patients) in the vax-plasma group and 6.2% (10 of 161 patients) in the standard of care group, which corresponded to a relative risk reduction of 65% (*P*=0.046). Evidence of efficacy in nonvaccinated patients cannot be inferred from these data because 94% (361 of 386 patients) of patients were vaccinated. In vaccinated patients with immunosuppression and COVID-19, the addition of vax-plasma or very high titer COVID-19 convalescent plasma to COVID-19 specific therapies reduced the risk of disease progression leading to hospitalization.

**IMPORTANCE:** As SARS-CoV-2 evolves, new variants of concern (VOCs) have emerged which evade available anti-spike monoclonal antibodies, particularly among immunosuppressed patients. However, high-titer COVID-19 convalescent plasma continues to be effective against VOCs because of its broad-spectrum immunomodulatory properties. Thus, we report clinical outcomes of 386 immunocompromised outpatients who were treated with COVID-19 specific therapeutics and a subgroup also treated with vaccine-boosted convalescent plasma. We found that administration of vaccine-boosted convalescent plasma was associated with a significantly decreased incidence of hospitalization among immunocompromised COVID-19 outpatients. Our data add to the contemporary data providing evidence to support the clinical utility of high-titer convalescent plasma as antibody replacement therapy in immunocompromised patients.

## Introduction

Although severe coronavirus disease 2019 (COVID-19) and hospitalization associated with COVID-19 are generally preventable among healthy vaccine recipients, patients with immunosuppression have poor immunogenic responses to COVID-19 vaccines and remain at high risk of infection with SARS-CoV-2 and hospitalization.^1,2^ Passive antibody therapy, via monoclonal antibody therapy or COVID-19 convalescent plasma, has been widely used to treat COVID-19, particularly among patients with immunosuppression.^3-5^ For example, in the outpatient setting, therapeutic use of neutralizing antispike monoclonal antibody has been associated with decreases in the incidence of COVID-19-related disease progression and hospitalization.^6^ However, monoclonal antibody therapy is limited by the emergence of novel SARS-CoV-2 variants that have serially escaped neutralization.^7,8^ Thus, although monoclonal antibody therapy as a cornerstone of COVID-19 treatment, at the time of this writing, there are no US FDA approved monoclonal antibodies for the treatment or prevention of SARS-CoV-2 infection.^6^ However, high-titer COVID-19 convalescent plasma continues to be effective against SARS-CoV-2 variants of concern (VOCs) because of its broad-spectrum immunomodulatory properties and ability to neutralize multiple SARS-CoV-2 variants.^9,10^ Although COVID-19 convalescent plasma is authorized for therapeutic use among patients with immunosuppression in the US and recommended by some organizations^11,12^, its use remains controversial.^4^

COVID-19 convalescent plasma collected from persons who have been both naturally infected with SARS-CoV-2 and vaccinated against SARS-CoV-2 (herein referred to as “vax-plasma” and also known as vaccine-boosted convalescent plasma) is particularly high titer, typically containing 10 to 100 times higher anti-SARS-CoV-2 antibody titers than standard COVID-19 convalescent plasma.^13-17^ To further our understanding of the clinical impact associated with vax-plasma, we report the clinical outcome of 386 immunocompromised outpatients who were diagnosed with COVID-19 and treated with contemporary COVID-19 specific therapeutics (standard of care group) and a subgroup who also received treatment with vax-plasma or high-titer COVID-19 convalescent plasma and (vax-plasma group) with a specific focus on hospitalization rates.

### Study design

This large, observational cohort study included data from a single health system (Mayo Clinic) and represented data from multiple health care sites across Minnesota and Wisconsin from 1 December 2022 to 1 December 2023. Immunocompromised patients with active COVID-19 infection, confirmed by SARS-CoV-2-specific reverse transcription polymerase chain reaction, were eligible to receive vax-plasma. The Mayo Clinic Institutional Review Board determined that this study met the criteria for exemption. Informed consent was waived. Only Mayo Clinic patients with research authorization were included.

As previously described^17^, eligible vax-plasma donors included individuals who had a confirmed diagnosis of COVID-19 and had received at least one dose of a SARS-CoV-2 vaccine. All donors experienced mild to moderate symptoms and met the national blood donor selection criteria. Vax-plasma was collected at least 10 days and up to 6 months after the complete resolution of COVID-19 symptomatology. Antibody titers of vax-plasma units met the minimum threshold required by the US FDA for high titer anti-SARS-CoV-2 antibodies, but precise antibody titers were not evaluated. However, numerous reports indicate that vax-plasma is uniformly extremely high titer with neutralizing activity against many SARS-CoV-2 variants^10,13,18^. The treatment schedule of vax-plasma transfusions was not standardized. Patients received the number of vax-plasma units deemed appropriate for each patient by their clinicians (range, 1 to 7 units).

The primary outcome was COVID-19–related hospitalization within 28 days after transfusion, assessed as the cumulative incidence in the vax-plasma group compared to the standard of care group who declined treatment vax-plasma. The decision to hospitalize patients was at the discretion of local clinicians. Continuous measures were compared between the treatment groups (vax-plasma group vs. standard of care group) using the two-sample t-test, whereas categorical measures were compared using the χ^2^ test or Fisher exact test, as appropriate. Reported p-values are two-sided and adjusted for multiplicity, as appropriate; and the interpretation of findings was based on p < 0.05.

## Results and discussion

Three-hundred eighty-six immunocompromised patients were offered standard of care treatments (e.g., remdesivir, nirmatrelvir or molnupiravir) and were also offered to be treated with vax-plasma. Of those patients, 58% agreed to treatment with vax-plasma (225 of 386 patients; vax-plasma group) and 42% (161 of 386 patients) received standard of care treatments alone without vax-plasma (standard of care group). Key demographic and clinical characteristics of the study population are provided in **Table 1**, stratified into the two treatment groups. Overall, the median age of all patients was 66 years (range: 2 to 96 years), 45% were female (175 of 386 patients), and 94% (361 of 386 patients) were vaccinated against SARS-CoV-2.

**Table 1.**
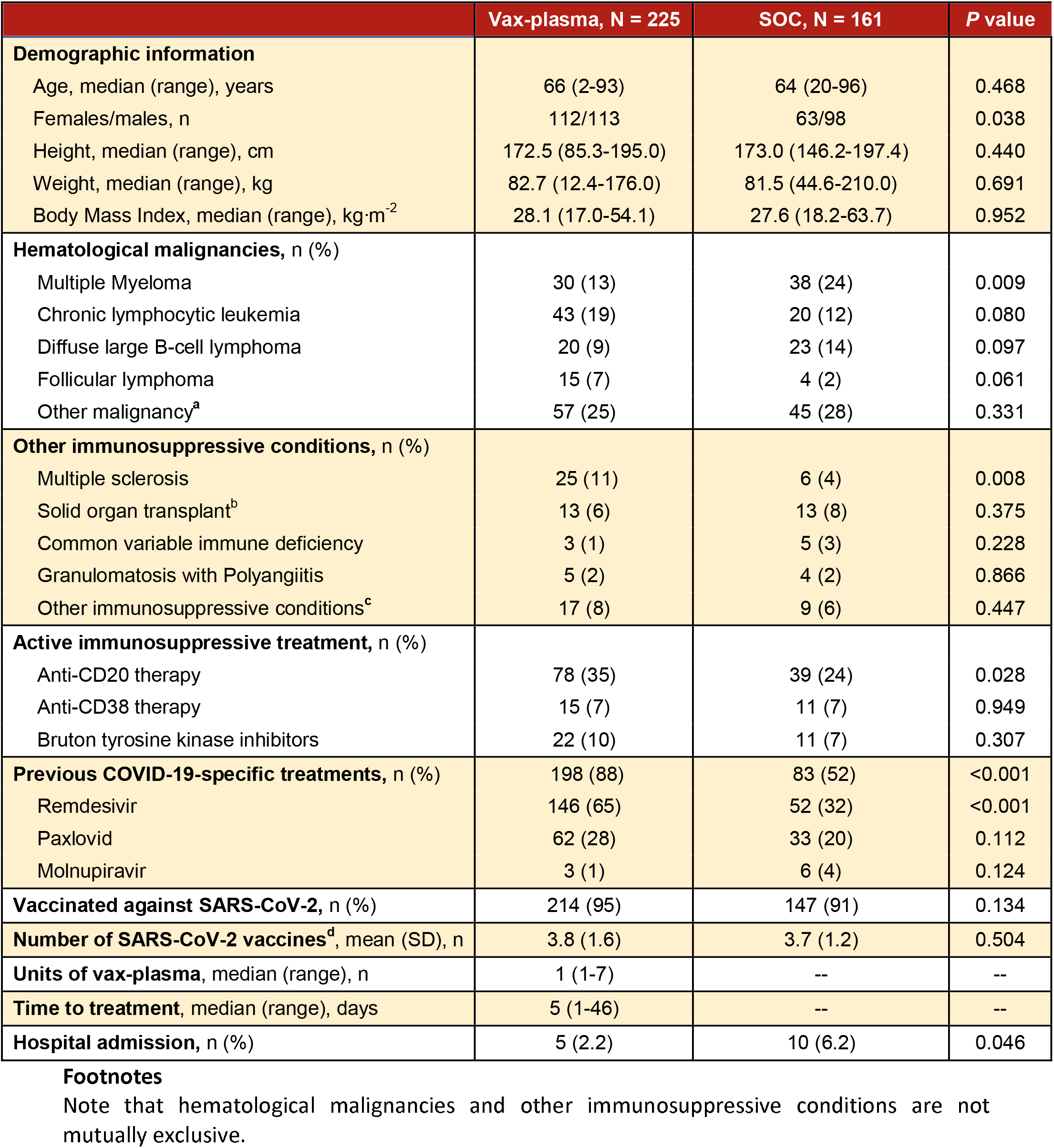
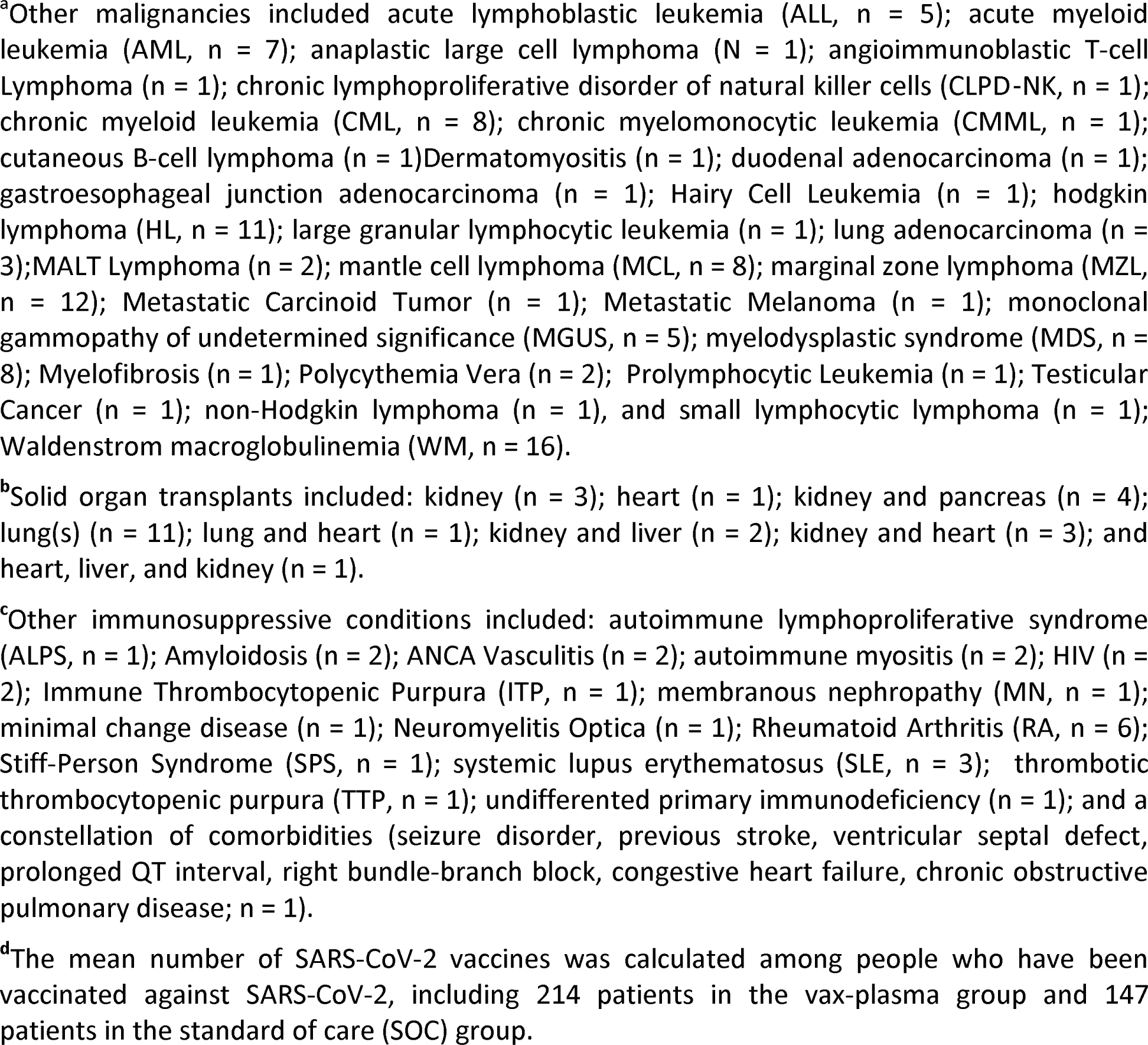
Characteristics of 386 immunocompromised outpatients who were diagnosed with COVID-19 and received standard of care COVID-19 therapeutics with or without vax-plasma.

Compared to patients in the standard of care group, patients in the vax-plasma group were more likely to be female (*P*=0.038), more likely to have received anti-CD20 monoclonal therapy (*P*=0.028), and more likely to have received previous COVID-19 antiviral treatments (*P*<0.001). Other key differences between the groups are noted in **Table 1**.

Patients had COVID-19 symptoms for a median of 5 days (range, 1 to 46 days) before receiving vax-plasma, and the median number of units transfused per patient was 1 (range, 1 to 7 units of vax-plasma). Most patients (73%, 281 of 386 patients) agreed to receive concomitant COVID-19–specific treatments, and patients in the vax-plasma group were more likely to have received COVID-19 specific treatments (88%, 198 of 225 patients) compared to the standard of care group (52%, 83 of 161 patients; *P*<0.001). COVID-19 specific treatments included remdesivir (65% of patients who accepted vax-plasma and 32% of those who declined vax-plasma), nirmatrelvir and ritonavir (PAXLOVID™) (28% in the vax-plasma group and 20% in the standard of care group), and/or molnupiravir (1% in the vax-plasma group and 4% in the standard of care group).

No major adverse effects were recorded among patients transfused with vax-plasma. The overall 28 day-hospital admission rate was 2.2% (5 of 225 patients) in vax-plasma group and 6.2% (10 of 161 patients) in standard of care group (*P*=0.046).

In this large, non-randomized cohort study involving outpatients with recent SARS-CoV-2 infection, the concomitant administration of high-titer COVID-19 convalescent plasma in addition to standard of care therapeutics was associated with a decreased incidence of hospitalization. Our observations are consistent with those of previous trials of antibody-based therapies —administration of sufficient pathogen-specific antibodies via COVID-19 convalescent plasma leads to a reduced risk of disease progression, COVID-19-related hospitalization, and COVID-19-related death in immunocompromised patients in both outpatient and inpatient settings.^5,19^ Collectively, contemporary clinical data provide evidence to support the utility of high-titer convalescent plasma including vax-plasma as antibody replacement therapy in immunocompromised patients.^17,20,21^

There is consensus that the primary mechanism of action of vax-plasma is through viral neutralization^19^—a finding established among humans early during the COVID-19 pandemic^22^ and supported by several preclinical models including mice,^23,24^ hamsters,^25^ and macaques.^26^ Because the neutralizing capacity of vax-plasma can evolve with emerging variants, issues related to escape by new variants that have time limited the efficacy of monoclonal antibodies can potentially be avoided. Additionally, sero-surveys of blood donors show a high prevalence of hybrid immunity in the population suggesting that very high titer vax-plasma is potentially available at a scale sufficient to treat immunocompromised patients.^27^ Thus, there is an emerging picture of utility for vax-plasma therapy in immunocompromised patients with SARS-CoV-2 infection that could benefit from further evaluation via carefully matched, larger real world data sets. Importantly, any prospective studies will need to consider the experimental design and ethical issues associated with potentially limiting a safe antibody therapy in immunocompromised patients unable to generate an adequate endogenous antibody response to infection.

Our study faced several contextual challenges associated with clinical research during a pandemic and limitations associated with the design of the study. First, the interpretation of these results is limited by the open-label and non-randomized design. In this framework, the vax-plasma group had higher rates of anti-CD20 monoclonal therapy, antiviral COVID-19-specific therapeutics, and vaccination against SARS-CoV-2. Importantly, treatment with anti-CD20 monoclonal therapy among people who are immunosuppressed is associated with the lowest likelihood to produce SARS-CoV-2 antibodies after one or more doses of COVID-19 vaccines.^16,28^ Thus, the effects of vax-plasma *per se* may not be definitively inferred. Second, the overall incidence of hospitalization was very low (3.8%, 15 of 386 patients), likely due in part to the very high number of patients who were vaccinated against SARS-CoV-2 and because most patients received standard of care COVID-19-specific therapeutics. In this context, the low incidence of the primary outcome (COVID-19–related hospitalization) limited the potential for definitive subgroup analyses according to coexisting immunosuppressive conditions or other putative confounding variables. Third, SARS-CoV-2 serology of vax-plasma units or patient samples was not systematically performed.

Despite the enumerated limitations of this study, our data provides evidence that transfusion of vax-plasma effectively transfers COVID-19-neutralizing antibodies to patients with immunosuppression and reduces the risk of COVID-19-related hospitalization. Vax-plasma appears to be an effective therapeutic throughout the clinical course of COVID-19 among immunocompromised patients from outpatients to inpatients with protracted COVID-19. For future pandemics, the use of therapeutic plasma with antibody levels in the upper deciles should be considered, particularly among immunocompromised patients.

## Acknowledgments

We thank the dedicated members of the Mayo Clinic Blood Donor Center for their rigorous efforts necessary to make this program possible. We also thank the donors who survived COVID-19 for providing vaccine-boosted COVID-19 convalescent plasma. Additionally, we thank Diana Zicklin Berrent and Chaim Lebovits for their progressive and passionate patient advocacy and support.

This work was supported by United Health Group and Mayo Clinic.

## Authorship Contributions

Study conception and design: J.G.R., S.M.T-S, M.J.J., and J.W.S. Acquisition, analysis, or interpretation of data: J.G.R., S.M.T-S, A.A.S., S.F., M.L.P., E.K.G., A.J.M., S.Z., E.A.O., and J.W.S. Drafting of the manuscript: J.G.R., S.M.T-S, M.J.J., and J.W.S. Administrative, technical, or material support: S.N.H., J.E.J., R.R.R., R.G., R.T.H., E.N.F., A.N.D., M.R.E., J.J.L., C.M.C., E.S.T., S.A.P., and N.E.K. All authors contributed to revising the manuscript, and all authors approved the final version of the manuscript.

## Data Availability

Datasets generated during this study may be available from corresponding authors on reasonable request. Requestors may be required to sign a data use agreement. Data sharing must be compliant with all applicable Mayo Clinic policies and those of the Mayo Clinic Institutional Review Board.

